# Toxoplasmosis: An important risk factor for acquiring SARS-CoV-2 infection and a severe course of Covid-19 disease

**DOI:** 10.1101/2021.05.15.21257257

**Authors:** Jaroslav Flegr

## Abstract

**Background:** Latent toxoplasmosis, i.e. a lifelong infection with the protozoan parasite *Toxoplasma* gondii, affects about a third of human population worldwide. In the past ten years, numerous studies had shown that infected subjects have a significantly higher incidence of mental and physical health problems and are more prone to exhibiting the adverse effects of various diseases.

**Methods:** A cross-sectional internet study was performed on a population of 4,499 *Toxoplasma*-free and 786 *Toxoplasma*-infected subjects and looked for factors which positively or negatively affect the risk of SARS-CoV-2 infection and likelihood of a severe course of Covid-19.

**Findings:** Logistic regression and partial Kendall correlation controlled for sex, age, and size of the place of residence showed that latent toxoplasmosis had the strongest effect on the risk of infection (OR = 1·50) before sport (OR = 1·30), and borreliosis (1·27). It also had the strongest effect on the risk of severe course of infection (Tau = 0·146), before autoimmunity, immunodeficiency, male sex, keeping a cat, being overweight, borreliosis, higher age, or chronic obstructive pulmonary disease. Toxoplasmosis augmented the adverse effects of other risk factors but was not the proximal cause of the effect of cat keeping (in the form of higher likelihood of Covid infection and higher severity of the course of infection), which was observed especially in a subset of *Toxoplasma*-infected subjects (Tau = 0·153). Effects of keeping a cat were detected only in subjects from multi-member families, suggesting that a cat could a vector for the transmission of SARS-CoV-2 within a family.

**Interpretations:** Toxoplasmosis is currently not considered a risk factor for Covid-19 and *Toxoplasma*- infected subjects are neither informed about their higher risks nor prioritised in vaccination programs. Because toxoplasmosis affects a large segment of the human population, its impact on Covid-19-associated effects on public health could be considerable.

## Introduction

About one-third of world’s population is infected by the protozoan parasite *Toxoplasma gondii*. The definitive host of *Toxoplasma* are cats (or any felines), but its intermediate host can be any warm-blooded animal, including humans. In immunocompetent human hosts, a short phase of acute toxoplasmosis spontaneously resolves into a latent phase. It is widely believed that during this phase, bradyzoites, the slowly reproducing stage of *Toxoplasma*, survive in tissue cysts localised mainly in immune-privileged organs and tissues for the duration of host’s life, waiting for the host to be eaten by a cat.^1^ For a long time, latent toxoplasmosis has been considered asymptomatic, which is why no effort was invested into finding a drug that would kill the bradyzoites in cysts and treat latent toxoplasmosis. In the past twenty years, however, numerous studies had shown that latent toxoplasmosis has important adverse effects on the mental and physical health of infected subjects.^2-4^ Over a hundred studies showed that latent toxoplasmosis strongly affects (according to meta-analytic studies nearly triples) the risk of schizophrenia. Later, a similar and possibly even stronger effect of toxoplasmosis was described in relation to other mental health disorders, including the obsessive-compulsive disorder, ADHD, and autism.^5, 6^

In the past decade, several studies showed that latent toxoplasmosis affects also physical health. The first evidence was merely indirect: researchers noticed that infected subjects show clear psychological symptoms of chronic stress, which explained most earlier observed differences in the personality profile and behaviour of *Toxoplasma*-infected versus *Toxoplasma*-free subjects.^7^ These changes, which go in the opposite direction in men and women^8, 9^ were at first interpreted as products of the parasite’s manipulative activity aimed at enhancing transmission from the intermediate host (usually a mouse) to the definitive host (a cat) by predation.^10^ A large cross-sectional study performed on 333 infected subjects and 1,153 controls showed that *Toxoplasma*- infected subjects reported higher rates of 77 from a list of 134 disorders.^11^ Infected subjects also scored significantly worse on 28 out of 29 health-related variables included in the study. Similar results were obtained in an ecological (correlational) study that was based on WHO data on the incidence of disease and disease burden in 88 WHO-member states.^12^ The results showed that the Disability Adjusted Life Year (DALY) for 23 out of 128 diseases and disease categories correlated with the prevalence of latent toxoplasmosis in individual countries even after controlling for per capita GDP, latitude, and humidity. In 29 European countries, differences in the prevalence of toxoplasmosis turned out to be responsible for 23% of total variability in disease burden in a model containing the three abovementioned covariates. The strongest associations were observed with cardiovascular diseases, perinatal conditions, and congenital abnormalities (which probably reflects the effect of congenital, not latent toxoplasmosis). Nevertheless, a strong positive association was observed also with filariasis, measles, and leukaemia. Such a broad range of observed associations suggests that the effect of latent toxoplasmosis is rather nonspecific. It has also been shown that toxoplasmosis makes individuals prone to various adverse factors, including fatigue, faster aging, and smoking.^13^ It is also known that *Toxoplasma* modifies the functioning of the host’s immune system, especially by increasing the concentration of certain lymphokines (most notably^14-17^ and changing the counts of various immunocytes.^18^

The main aim of the present study was to explore whether latent toxoplasmosis has any effect on the risk of SARS-CoV-2 infection and the course of Covid-19 disease. To this purpose, the probability of being diagnosed with Covid-19, severity of the course of the disease, and the incidence of specific symptoms of the disease were compared between 481 *Toxoplasma*-infected and 4,018 *Toxoplasma*- free participants of a large internet survey.

## Material and Methods

Participants were recruited by a Facebook-based snowball method.^19^ Calls for participation in the study were published about 15 times on the Facebook page of Labbunnies, a 23,000-member group of Czech and Slovak nationals willing to take part in evolutionary psychology studies and help with recruiting participants of such studies, as well as on the authors’ personal Facebook and Twitter accounts. The Qualtrics questionnaire that was used to gather data contained Facebook ‘share’ and ‘like’ buttons, so that participants could help recruit other participants by pressing these buttons. In total, the buttons were pressed 12,000 times between 17 October 2020 and 3 March 2021. In total, data from 52,000 respondents were obrtained. The invitation as well as the informed consent on the first page of the questionnaire contained only most general information about the aims of the study and contents of the questionnaire. The subjects were informed that the study examines which factors affect the risk and course of Covid-19 infection and what is the opinion of people about anti-epidemic measurements. Participants were also informed that their participation is voluntary, that they can skip any questions they might find uncomfortable and terminate their cooperation at any point simply by closing the web page. Only subjects who consented to participate in the study by pressing the corresponding button were allowed to take the questionnaire. Respondents were not paid for their participation in the study, but after finishing the 20-minute questionnaire, they received information about the results of related studies. The study was anonymous, but participants had the option of providing their e-mail addresses for a future longitudinal study (about 42% did) or could ask for their data to be erased after completing the questionnaire (about 2% did). Data collection was performed in accordance with the relevant guidelines and regulations and the project, including the method of obtaining from all participants informed consent with participation in this anonymous study, was approved by the Institutional Review Board of the Faculty of Science, Charles University (Komise pro práci s lidmi a lidským materiálem Přírodovědecké Fakulty Univerzity Karlovy) — No. 2020/25. The study was preregistered at The Open Science Framework: https://doi.org/10.17605/OSF.IO/VWXJE

### Questionnaire

The Qualtrics survey consisted of three parts related to three different projects (Risk and protective factors, Opinions of the Czech public on the anti-epidemic measurements, and the effect of priming by studying graphs of Covid victims on opinions regarding anti-epidemic measures). In the present study, only responses to questions related to Covid-19 risks and protective factors were inspected and analysed. Respondents were asked about their sex, age, size of their place of residence (scale 1– 5, 0: under 1,000 inhabitants, 1: 1–5,000 inhabitants, 2: 5–50,000 inhabitants, 3: 50–100,000 inhabitants, 4: 100–500,000 inhabitants, 5: over 500,000 inhabitants), and how many persons live with them in the same household. The subjects indicated whether they had already contracted Covid-19 by choosing from five answers (1: ‘No’, 2: ‘Yes, I was diagnosed with it’, 3: ‘Yes, but I was not diagnosed with it’, 4: ‘I am awaiting test results’, 5: ‘No, but I was in a quarantine’). For the purpose of the current study, answers 1 and 5 were coded as 0 (uninfected with Covid), answer 2 as 1 (Covid patients), and answers 3 and 4 were coded as NA (data not available). The respondents were also asked whether they had ever been tested in a laboratory for toxoplasmosis and/or borreliosis, and what the result of these tests was (negative/positive-infected/’I do not know, I am not sure’). For both toxoplasmosis and borreliosis, the questionnaire was pre-set to indicate the third response ‘I do not know, I am not sure’ as a default. Similarly, respondents were asked about their Rh status (positive/negative/’I do not know, I am not sure’) and blood group (A/B/AB/0/’I do not know, I am not sure’). Respondents were also asked to indicate which risks and protective factors apply to them (for a list of the corresponding binary variables, see column 1 of Table 1). Those who had been diagnosed with Covid-19 were also asked to rate the severity of the course of the disease on a five-point scale (1: no symptoms, 2: like a mild flu, 3: like a severe flu, 4: I was hospitalised, 5: I was treated at an ICU). They also had to check which symptoms they experienced during the course of the Covid infection (for a list of the corresponding binary variables, see column 1 of Table 3).

**Table 1.** Incidence of risk and protective factors for Covid-19 in men and women

|  | Women |  |  |  | Men |  |  |  | OR | C.I.<sub>.95</sub> | p |
| --- | --- | --- | --- | --- | --- | --- | --- | --- | --- | --- | --- |
|  | no Covid |  | Covid |  | no Covid |  | Covid |  |  |  |  |
|  | N | % | N | % | N | % | N | % |  |  |  |
| Toxoplasmosis | 563 | 18.31 | 81 | 23.89 | 115 | 12.05 | 27 | 20.61 | **1.50** | **1.19–1.89** | **0.0007** |
| Borreliosis | 410 | 19.85 | 57 | 24.26 | 108 | 14.57 | 17 | 17.00 | 1.27 | 0.96–1.68 | 0.0881 |
| Rh-positivity | 1953 | 76.41 | 219 | 76.31 | 490 | 75.85 | 61 | 74.39 | 0.98 | 0.76–1.26 | 0.8854 |
| A blood group | 989 | 37.26 | 123 | 41.14 | 236 | 34.05 | 33 | 35.87 | 1.16 | 0.93–1.44 | 0.1795 |
| B blood group | 568 | 21.40 | 50 | 16.72 | 156 | 22.51 | 22 | 23.91 | 0.82 | 0.62–1.07 | 0.1368 |
| AB blood group | 247 | 9.31 | 34 | 11.37 | 91 | 13.13 | 13 | 14.13 | 1.19 | 0.86–1.65 | 0.2929 |
| 0 blood group | 850 | 32.03 | 92 | 30.77 | 210 | 30.30 | 24 | 26.09 | 0.91 | 0.73–1.15 | 0.4396 |
| Being overweight | 739 | 35.11 | 90 | 31.03 | 258 | 42.64 | 35 | 33.02 | 0.74 | 0.57–0.95 | 0.0169 |
| Diabetes | 75 | 3.56 | 7 | 2.41 | 28 | 4.63 | 8 | 7.55 | 0.96 | 0.52–1.76 | 0.8921 |
| Cardiovascular problems | 131 | 6.22 | 16 | 5.52 | 65 | 10.74 | 9 | 8.49 | 0.81 | 0.50–1.30 | 0.3820 |
| Asthma | 1846 | 87.70 | 256 | 88.28 | 544 | 89.92 | 89 | 83.96 | 1.13 | 0.80–1.59 | 0.4908 |
| Chronic obstructive pulmonary disease | 49 | 2.33 | 5 | 1.72 | 16 | 2.64 | 2 | 1.89 | 0.67 | 0.27–1.63 | 0.3737 |
| Immunosuppression | 289 | 13.73 | 40 | 13.79 | 52 | 8.60 | 9 | 8.49 | 1.00 | 0.70–1.42 | 0.9929 |
| Allergy | 548 | 26.03 | 77 | 26.55 | 150 | 24.79 | 28 | 26.42 | 1.06 | 0.81–1.37 | 0.6798 |
| Autoimmunity | 168 | 14.35 | 25 | 11.57 | 26 | 8.72 | 6 | 9.09 | 0.75 | 0.45–1.24 | 0.2545 |
| Living alone | 285 | 9.30 | 28 | 8.28 | 160 | 16.91 | 23 | 17.56 | 0.96 | 0.70–1.32 | 0.8027 |
| Tobacco smoking | 415 | 19.71 | 54 | 18.62 | 148 | 24.46 | 23 | 21.70 | 0.89 | 0.66–1.19 | 0.4341 |
| Marihuana consumption | 56 | 2.66 | 6 | 2.07 | 40 | 6.61 | 14 | 13.21 | 1.49 | 0.88–2.53 | 0.1367 |
| Daily alcohol consumption | 126 | 5.99 | 16 | 5.52 | 97 | 16.03 | 14 | 13.21 | 0.86 | 0.55–1.33 | 0.4887 |
| Frequent singing | 269 | 12.78 | 39 | 13.45 | 50 | 8.26 | 14 | 13.21 | 1.21 | 0.87–1.70 | 0.2612 |
| Sport | 620 | 29.45 | 92 | 31.72 | 274 | 45.29 | 60 | 56.60 | 1.30 | 1.03–1.65 | 0.0300 |
| Cold water swimming | 205 | 9.74 | 37 | 12.76 | 102 | 16.86 | 19 | 17.92 | 1.32 | 0.95–1.84 | 0.0989 |
| Frequent sauna use | 94 | 8.03 | 20 | 9.26 | 37 | 12.42 | 8 | 12.12 | 1.16 | 0.70–1.93 | 0.5558 |
| Vitamins and supplements | 753 | 35.77 | 133 | 45.86 | 289 | 47.77 | 60 | 56.60 | **0.62** | **0.49–0.78** | **0.0001** |
| Wearing glasses | 1101 | 60.36 | 169 | 61.90 | 371 | 67.82 | 67 | 66.34 | 0.96 | 0.75–1.24 | 0.7723 |
| Dog keeping | 365 | 44.19 | 78 | 49.37 | 93 | 51.10 | 21 | 45.65 | 0.82 | 0.56–1.18 | 0.282 |
| Cat keeping | 381 | 46.13 | 82 | 51.90 | 82 | 45.05 | 17 | 36.96 | 1.16 | 0.80–1.68 | 0.4303 |

*The last four columns show the results of logistic regression with Covid-19 infection as a dependent variable, age, sex, size of the place of residence as covariates, and one of risk/protective factors listed in the first column as the independent variable. Results shown in bold were significant after correction for multiple tests.*

### Statistical analyses

Before embarking on any analyses, all subjects who completed the questionnaire for the second or third time (they were asked about this at the end of the questionnaire for purposes of the planned longitudinal study), those who answered all or nearly all questions by the same code, and those who finished the questionnaire in less than six minutes were filtered out. After this filtration, the set contained 29,345 records. Only the subjects who were negatively or positively tested for toxoplasmosis (4,499 subjects) were included in the present study.

Statistical analysis was performed with the Statistica v. 10.0. (descriptive statistics, t-tests) and R v. 3.3.1^20^ (all other tests) packages. To compute the partial Kendall correlation, the ppcor package was used.21 Correction for multiple tests was done using Benjamini-Hochberg procedure with false discovery rate pre-set to 0·10.^22^ The datasets generated and/or analysed during the current study are available in Figshare repository, 10.6084/m9.figshare.14559993.

## Results

The final set contained 1,085 men, 142 (13·09%) of whom were infected with *Toxoplasma* and 131 (12·07%) diagnosed with Covid, and 3,414 women, 644 (18·86%) of whom were infected with *Toxoplasma* and 339 (9·93%) diagnosed with Covid. The incidence of other potential risk factors shows Table 1.

A significant difference in age between men (38·9, s.d. 10·9) and women (40·0, s.d. 10·5) (t_4497_ = - 2·86, p = 0·0043) and between *Toxoplasma*-free (39·3, s.d. 10·5) and *Toxoplasma*-infected (42·7, s.d. 10·1) women (t_3412_ = -7·41, p < 0·0001), but not between *Toxoplasma*-free (38·7, s.d. 10·7) and *Toxoplasma*-infected (40·0, s.d. 11·8) men (t_1083_ = -1·25, p = 0·211) existed in the population under study. No association between age and Covid in either men or women (p > 0·38) was detected.

The risk of acquiring toxoplasmosis depends on certain confounding factors, such as sex, age, and size of the place of residence. Therefore, the association between potential risk factors and protective factors was analysed using a logistic regression with binary variable Covid as a dependent variable, sex, age, size of place of residence as covariates, and one of the analysed factors as the independent variable. The results are shown in the last three columns of Table 1. Toxoplasmosis, with an odds ratio of 1·50 (C.I._95_ = 1·19–1·89, p = 0·0007, risk ratio 1·40 (CI_95_: 1·05–1·80) for women and 1·91 (CI_95_: 1·20–3·05) for men, turned out to be the most serious risk factor for being diagnosed with Covid.

It is known that Rh-negative subjects are more prone to experiencing the negative effects of toxoplasmosis than Rh-positive subjects are.23-25 Therefore, also a more complex model containing not only sex, age, size of the place of residence, and toxoplasmosis, but also the Rh factor and Rh– toxoplasmosis interaction was analysed. Logistic regression showed a significant effect of toxoplasmosis (OR = 1·45, CI_95_: 1·06–2·10, p = 0·046), but not Rh (p = 0·86) or Rh–toxoplasmosis interaction (p = 0·84).

Respondents who had been diagnosed with Covid-19 were asked to rate the course of their disease and indicate which symptoms they experienced during their illness. Figure 1 shows that *Toxoplasma*- infected subjects of both sexes had a more serious course of Covid. Partial Kendall correlation (Table 2) showed that toxoplasmosis represents a more pronounced risk factor for a severe course of Covid-19 than compromised autoimmunity, immunodeficiency, male sex, cat keeping, being overweight, borreliosis, higher age, or chronic obstructive pulmonary disease does.

**Fig. 1.**
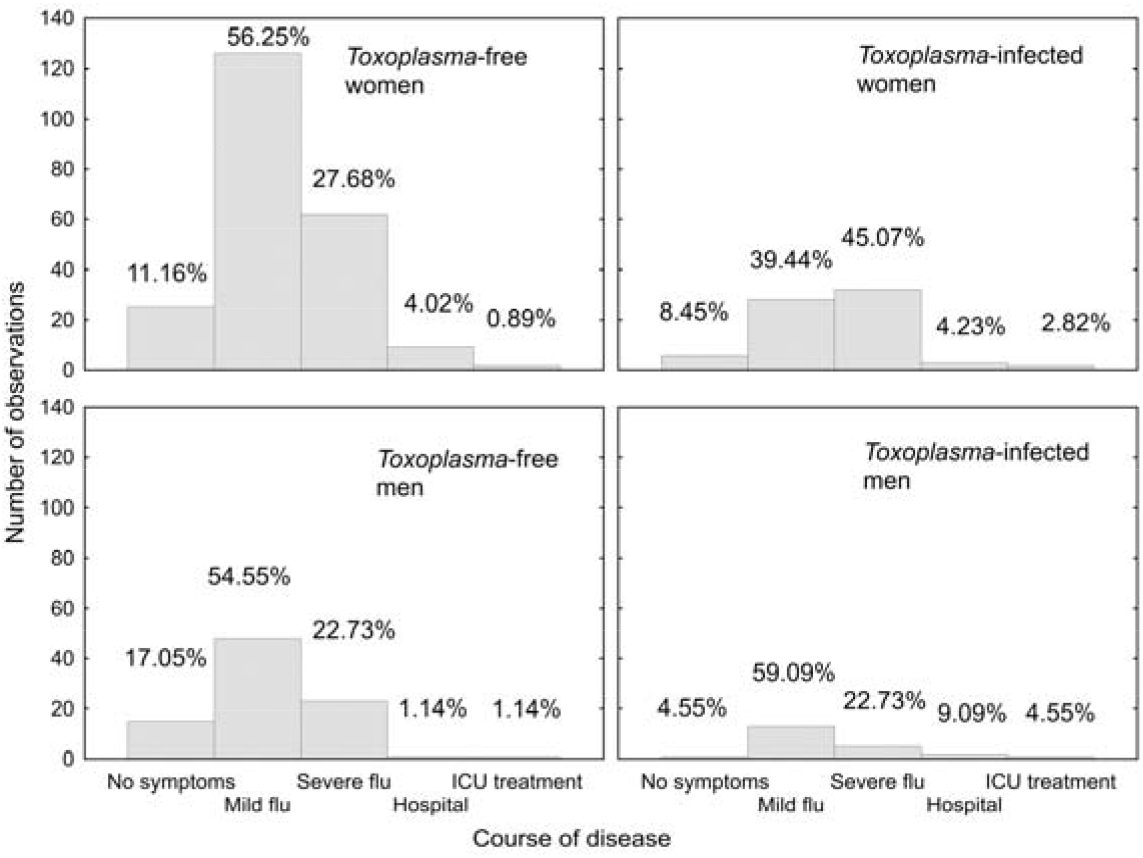
The course of Covid-19 in *Toxoplasma*-free and *Toxoplasma*-infected men and women

**Table 2.** Correlation between various risk/protective factors and severity of Covid-19 disease.

|  | All |  | Women |  | Men |  | Toxo-free |  | Toxo-infected |  |
| --- | --- | --- | --- | --- | --- | --- | --- | --- | --- | --- |
|  | Tau | p | Tau | p | Tau | p | Tau | p | Tau | p |
| Sex | **0.116** | **0.0002** | NA | NA | NA | NA | **0.112** | **0.0017** | 0.095 | 0.1713 |
| Age | **0.093** | **0.0032** | **0.124** | **0.0010** | 0.025 | 0.6763 | 0.066 | 0.0654 | 0.137 | 0.0494 |
| Size of place of residence | 0.009 | 0.7815 | -0.002 | 0.9575 | 0.039 | 0.5093 | 0.000 | 0.9895 | 0.065 | 0.3477 |
| Toxoplasmosis | **0.146** | **0.0000** | **0.150** | **0.0001** | 0.123 | 0.0376 | NA | NA | NA | NA |
| Borreliosis | **0.094** | **0.0123** | **0.120** | **0.0088** | 0.030 | 0.6529 | 0.010 | 0.8146 | 0.212 | 0.0301 |
| Rh-positivity | 0.036 | 0.3097 | 0.035 | 0.3929 | 0.064 | 0.3950 | 0.022 | 0.5941 | 0.098 | 0.2112 |
| A blood group | 0.019 | 0.5892 | 0.000 | 0.9945 | 0.079 | 0.2715 | 0.032 | 0.4210 | -0.052 | 0.4932 |
| B blood group | -0.008 | 0.8101 | -0.010 | 0.7940 | 0.009 | 0.8991 | 0.012 | 0.7680 | -0.069 | 0.3606 |
| AB blood group | 0.021 | 0.5371 | **0.093** | **0.0210** | -0.160 | 0.0254 | -0.002 | 0.9586 | 0.080 | 0.2909 |
| O blood group | -0.028 | 0.4278 | -0.050 | 0.2142 | 0.031 | 0.6694 | -0.043 | 0.2834 | 0.048 | 0.5260 |
| Being overweight | **0.104** | **0.0012** | **0.107** | **0.0052** | 0.116 | 0.0558 | 0.089 | 0.0147 | 0.140 | 0.0505 |
| Diabetes | 0.004 | 0.8922 | -0.021 | 0.5777 | 0.038 | 0.5301 | 0.019 | 0.5929 | -0.022 | 0.7602 |
| Cardiovascular problems | 0.063 | 0.0514 | 0.063 | 0.0985 | 0.075 | 0.2194 | 0.049 | 0.1762 | 0.170 | 0.0174 |
| Asthma | 0.052 | 0.1055 | 0.036 | 0.3496 | 0.094 | 0.1233 | 0.039 | 0.2815 | 0.133 | 0.0630 |
| Chronic obstructive pulmonary disease | **0.072** | **0.0252** | 0.073 | 0.0583 | 0.080 | 0.1853 | 0.013 | 0.7220 | **0.219** | **0.0022** |
| Immunodeficiency | **0.135** | **0.0000** | **0.141** | **0.0002** | 0.116 | 0.0558 | **0.160** | **0.0000** | 0.067 | 0.3472 |
| Allergy | -0.015 | 0.6453 | -0.011 | 0.7829 | -0.025 | 0.6809 | -0.028 | 0.4376 | 0.025 | 0.7235 |
| Autoimmunity | **0.137** | **0.0004** | **0.146** | **0.0011** | 0.112 | 0.1590 | 0.074 | 0.0874 | **0.254** | **0.0048** |
| Living alone | 0.034 | 0.2794 | **0.092** | **0.0151** | -0.061 | 0.2999 | 0.030 | 0.3960 | 0.073 | 0.2990 |
| Tobacco smoking | 0.061 | 0.0576 | 0.045 | 0.2454 | 0.090 | 0.1364 | 0.082 | 0.0239 | 0.006 | 0.9351 |
| Marihuana consumption | -0.001 | 0.9777 | **-0.082** | **0.0330** | 0.081 | 0.1800 | 0.057 | 0.1207 | **-0.195** | **0.0065** |
| Daily alcohol consumption | -0.039 | 0.2311 | **-0.094** | **0.0138** | 0.067 | 0.2677 | -0.055 | 0.1279 | 0.035 | 0.6289 |
| Frequent singing | -0.032 | 0.3162 | -0.060 | 0.1153 | 0.056 | 0.3553 | -0.053 | 0.1459 | -0.001 | 0.9861 |
| Sport | **-0.092** | **0.0044** | **-0.112** | **0.0036** | -0.056 | 0.3543 | **-0.116** | **0.0015** | -0.028 | 0.6944 |
| Cold water swimming | -0.047 | 0.1447 | -0.041 | 0.2851 | -0.055 | 0.3614 | -0.062 | 0.0883 | -0.041 | 0.5691 |
| Frequent use of sauna | -0.036 | 0.3550 | -0.040 | 0.3731 | -0.037 | 0.6432 | -0.009 | 0.8280 | -0.177 | 0.0491 |
| Vitamins and supplements | -0.047 | 0.1425 | -0.029 | 0.4437 | -0.087 | 0.1499 | -0.068 | 0.0628 | 0.034 | 0.6329 |
| Wearing glasses | -0.033 | 0.3244 | 0.005 | 0.9037 | -0.131 | 0.0346 | -0.003 | 0.9346 | -0.069 | 0.3559 |
| Current dog keeping | 0.004 | 0.9321 | 0.015 | 0.7737 | -0.042 | 0.6694 | 0.049 | 0.3497 | -0.120 | 0.2637 |
| Current cat keeping | **0.107** | **0.0203** | **0.123** | **0.0194** | 0.058 | 0.5549 | 0.084 | 0.1052 | 0.153 | 0.1544 |

*The table shows effect size and significance, i.e. partial Kendall Taus and p values. The effect of sex was controlled for age, and size of place of residence, the effect of age for sex, and size of place of residence, and size of place of residence for sex and age; all other effects were controlled for all three covariates. Positive Tau means a more severe course of Covid in subjects reporting the factor listed in the first column. The results in bold were significant after correction for multiple tests. In less numerous men, all factors lost their significance after correction for multiple tests but observed Tau values show that the effects in men are mostly stronger than in women.*

**Table 3.** Symptoms of Covid-19 in *Toxoplasma*-infected and *Toxoplasma*-free subjects

|  | All |  | Women |  | Men |  | OR | C.I.<sub>95</sub> | p | OR<sub>W</sub> | OR<sub>M</sub> |
| --- | --- | --- | --- | --- | --- | --- | --- | --- | --- | --- | --- |
|  | Toxo | Toxo | Toxo | Toxo | Toxo | Toxo |  |  |  |  |  |
|  | - | + | - | + | - | + |  |  |  |  |  |
| Fever >38°C (100.4 °F) | 0.38 | 0.39 | 0.34 | 0.37 | 0.47 | 0.45 | 1.07 | 0.67–1.7 | 0.789 | 1.09 | 0.99 |
| Fatigue | 0.74 | 0.81 | 0.79 | 0.90 | 0.57 | 0.57 | 1.19 | 0.45–3.14 | 0.729 | 1.30 | 1.22 |
| Dry cough | 0.42 | 0.53 | 0.44 | 0.50 | 0.38 | 0.64 | 1.40 | 0.89–2.2 | 0.141 | 1.16 | 2.97 |
| Shortness of breath | 0.32 | 0.39 | 0.33 | 0.41 | 0.31 | 0.32 | 1.19 | 0.74–1.91 | 0.466 | 1.30 | 1.04 |
| Sore throat | 0.36 | 0.36 | 0.37 | 0.40 | 0.34 | 0.23 | 1.00 | 0.63–1.61 | 0.985 | 1.08 | 1.36 |
| Headache | 0.70 | 0.78 | 0.74 | 0.84 | 0.60 | 0.59 | 1.37 | 0.82–2.29 | 0.225 | 1.71 | 0.90 |
| Chest pain or pressure | 0.32 | 0.51 | 0.35 | 0.53 | 0.24 | 0.45 | **1.96** | **1.24–3.13** | **0.004** | 1.79 | 2.73 |
| Other pains | 0.31 | 0.38 | 0.33 | 0.41 | 0.26 | 0.27 | 1.20 | 0.74–1.93 | 0.453 | 1.16 | 1.18 |
| Diarrhoea | 0.28 | 0.36 | 0.29 | 0.36 | 0.25 | 0.36 | 1.40 | 0.86–2.26 | 0.171 | 1.17 | 2.26 |
| Conjunctivitis | 0.08 | 0.12 | 0.09 | 0.14 | 0.05 | 0.05 | 1.28 | 0.6–2.72 | 0.522 | 1.22 | 1.84 |
| Loss of smell | 0.71 | 0.72 | 0.73 | 0.74 | 0.66 | 0.64 | 0.99 | 0.61–1.59 | 0.954 | 0.99 | 0.94 |
| Loss of taste | 0.58 | 0.55 | 0.62 | 0.59 | 0.49 | 0.45 | 0.88 | 0.56–1.39 | 0.593 | 0.89 | 0.87 |
| Middle ear pain | 0.12 | 0.22 | 0.14 | 0.25 | 0.06 | 0.14 | 1.95 | 0.68–5.56 | 0.209 | 1.67 | 5.11 |
| Problems speaking and walking | 0.03 | 0.05 | 0.03 | 0.06 | 0.04 | 0.05 | 1.72 | 0.6–4.98 | 0.313 | 1.32 | 2.18 |
| Skin rash | 0.08 | 0.10 | 0.10 | 0.09 | 0.02 | 0.14 | 1.31 | 0.6–2.87 | 0.492 | 0.77 | 6.00 |
| Changes in pigmentation | 0.03 | 0.05 | 0.04 | 0.06 | 0.01 | 0.05 | 1.81 | 0.64–5.09 | 0.262 | 1.29 | 4.05 |
| Higher sexual appetite | 0.03 | 0.08 | 0.03 | 0.06 | 0.04 | 0.14 | **3.40** | **1.42–8.17** | **0.006** | 2.53 | 4.50 |
| Other symptoms | 0.27 | 0.25 | 0.29 | 0.27 | 0.22 | 0.18 | 0.93 | 0.56–1.55 | 0.778 | 0.90 | 0.99 |

*Columns 2–7 show reported incidence of particular symptoms (in percents); the last five columns show results logistic regression with a particular symptom as the dependent variable, age, sex, size of place of residence as covariates, and toxoplasmosis as the independent variable. The last two columns show OR computed separately for women and men. The results in bold were significant after correction for multiple tests.*

One could speculate that keeping a cat could be just a proxy for being *Toxoplasma*-infected and indeed, the prevalence of toxoplasmosis in subjects who do keep a cat (24.6%) was markedly higher than in those who do not (15·5%) (Chi^2^ = 15·5, p<0.0001). To test this hypothesis, the partial Kendall correlation tests was performed separately for *Toxoplasma*-free and *Toxoplasma*-infected subjects (Table 2, the last four columns). Existence of the cat-keeping effect (i.e. increased likelihood of contracting Covid-19 and of having a severe course of the disease) in the *Toxoplasma*-infected subpopulation suggests that keeping a cat is a real risk factor for Covid-19, not just a side-effect of a higher probability of having toxoplasmosis. In fact, the strength of the cat-keeping effect was twice higher in the *Toxoplasma*-infected (Tau = 0·153) than in *Toxoplasma*-free subjects (Tau = 0·084). One could speculate that a cat might be able to transmit Covid among family members. Members of one family could moreover be infected repeatedly and the virus could adapt to the similar genotype of (genetically related) members of the same family; both of the above scenarios could then result in a more severe course of the disease. Indeed, separate analyses for 194 respondents who lived in multi-member families showed that keeping a cat increases the risk of a severe course of Covid-19 (Tau = 0·116, p = 0·017). The same analysis for 22 respondents who lived alone showed no such increase; in fact, it showed a nonsignificant effect in the opposite direction (Tau = -0·028, p = 0·867). Analogical analyses of the risk of acquiring SARS-CoV-2 infection showed no significant effect of cat keeping neither in people who live in multi-member families nor in those who live alone. However, even here a positive effect of cat keeping in 3,983 respondents living in multi-member families (OR = 1·210, CI_95_: 0.82–1·78, p = 0·332) and a negative effect in 496 respondents living alone (OR = 0·692, CI_95_: 0·20– 2·46, p = 0·568) were found.

The results also showed that many factors taken into consideration in this study had a much stronger effect on the risk of severe course of Covid in *Toxoplasma*-infected subjects than in those who were *Toxoplasma*-free: borreliosis (Tau = 0·212 vs. 0·010), being overweight (Tau = 0·140 vs. 0·089), cardiovascular diseases (Tau = 0·170 vs. 0·049), asthma (Tau = 0·133 vs. 0·039), chronic obstructive pulmonary disease (Tau = 0·219 vs. 0·013), allergy (Tau = 0·089 vs. 0·012), compromised autoimmunity (Tau = 0·254 vs. 0·074), and cat keeping (Tau = 0·153 vs. 0·0840). Conspicuous exceptions were male sex (Tau = 0·095 vs. 0·112) and immunodeficiency (Tau = 0·067 vs. 0·160). The protective effect of sport was also stronger in the *Toxoplasma*-free than in the *Toxoplasma*-infected subset (Tau = -1·16 vs. 0·028).

Logistic regression controlled for age, sex, and size of the place of residence showed that *Toxoplasma*-infected participants reported having more symptoms during Covid, but only the presence of chest pain or pressure (OR = 1·96) and higher sexual appetence (OR = 2·68) were statistically significant after correction for multiple tests (Table 3). On the other hand, even the effects that did not reach the formal level of statistical significance tended to show a higher frequency of symptoms in *Toxoplasma*-infected than in *Toxoplasma*-free subjects and were nearly always stronger in men than in women.

## Discussion

*Toxoplasma*-infected subjects had a higher probability of being diagnosed with Covid-19 and of having a more severe course of the disease; they were more likely to end up hospitalised and more frequently needed to be treated at intensive care units. They more frequently expressed the specific Covid symptoms, especially the serious ones such as difficulties with speaking and walking.

Interestingly, they also more frequently reported higher sexual appetence during the disease. This symptom was reported by 24·0% (6 out of 25) *Toxoplasma*-infected and 6·45% (6 out of 93) *Toxoplasma*-free male patients with Covid and in 6·41% (5 out of 78) *Toxoplasma*-infected and 2·5% (6 out of 238) *Toxoplasma*-free female patients with Covid. Originally, this trait was included on the list of symptoms because higher sexual appetence was reported to be related to transition from a slower to faster life strategy in subjects who are in worse health.26 In agreement with this expectation, it was reported more often by subjects who had a more serious course of the disease, and specifically in *Toxoplasma*-infected male patients. On the other hand, it is unclear whether higher sexual appetence is related to Covid or just their *Toxoplasma* infection: numerous independent studies had shown that *Toxoplasma*-infected males (both rodents and men) have higher testosterone levels27-32 and higher concentration of this steroid hormone could indeed be the proximal cause of their higher sexual appetence. It will be necessary to collect data on the difference in sexual appetence of *Toxoplasma*-infected and *Toxoplasma*-free men without Covid in future studies.

The effects of toxoplasmosis on the risk of Covid and risk of severe course of Covid-19 were the strongest effects detected in the present study. It must be, however, remembered that subjects with various known risk factors, such as being overweight, are probably more careful and actively try to avoid possible sources of the infection. It important to note that most factors known to be linted to a severe course of Covid decreased (albeit usually non-significantly) the risk of becoming infected. *Toxoplasma* infection is not, however, considered a factor that could increase the risk of severe course of Covid and *Toxoplasma*-infected persons therefore do not modify their behaviour so as to minimise the risk of becoming infected with Covid. Consumption of vitamins and food supplements seems to have the strongest protective effect on the risk of becoming infected with Covid, but people who take vitamins probably also apply other measures to avoid the infection, which is why nobody can tell whether vitamins and supplements alone have such a strong protective effect. Sport had the strongest protective effect against a severe course of Covid-19, but it is also a strong risk factor for acquiring the infection, probably because sport activities increase the number of contacts with potential sources of SARS-CoV-2 infection.

Our data showed that latent toxoplasmosis was the strongest risk factor for a severe course of Covid-19. It was stronger than the effect of being overweight, cardiovascular disease, or diabetes. The relative strength of the particular effects should be, however, interpreted with caution. It is possible that subjects with the most severe course of Covid (and logically also those who died) did not participate in our questionnaire study. This preselection could probably explain the relatively weak effect of chronic obstructive pulmonary disease.

There has been much speculation as well as some indications that cats could be a vector of Covid-19 and transmit the virus at least within a family.33-35 Our data seem to support this possibility. The nonsignificant positive effect of cat keeping was present in respondents from multimember families, and negative, though again nonsignificant, effect of cat keeping was observed in respondents who live alone. Unexpectedly, keeping a cat was found to increases the risk of a severe course of Covid: it was the fifth strongest factor after toxoplasmosis, diagnosed immunodeficiency, autoimmunity, and male sex. The existence of this effect in a subpopulation of *Toxoplasma*-infected subjects invalidates the hypothesis that cat keeping is only a proxy of *Toxoplasma* infection (and possibly a better proxy than the presence of anti-IgG antibodies). The negative effect of cat keeping on health is in line with previously published data. It has been shown that people who keep a cat are significantly more likely to develop many mental disorders, including the bipolar disorder (OR = 2·66)^36^. Much stronger and more numerous associations were observed between mental health and being bitten^37^ or scratched^38, 39^ by a cat. It has even been suggested that the cat scratch disease, i.e. infection by bacteria from the genus Bartonella^36^ or another unknown pathogen^40^, could be responsible for the effects of the cat-related injuries on the mental and physical health of subjects who keep a cat.

Analyses performed separately on the *Toxoplasma*-free and *Toxoplasma*-infected subjects had also shown that most factors have a much stronger effect (or exist only) in the *Toxoplasma*-infected subpopulation. For example, the effect of borreliosis is 21·5, chronic obstructive pulmonary disease 16·8, cardiovascular diseases 3·5, compromised autoimmunity 3·4, and age 2·1-times stronger in the *Toxoplasma*-infected than in *Toxoplasma*-free subjects. This enhancing effect of toxoplasmosis applies also to some protective factors: visits to sauna and marihuana consumption protect *Toxoplasma*-infected but not *Toxoplasma*-free subjects from infection. In contrast, the protective effect of sport was about four times stronger in *Toxoplasma*-free than in *Toxoplasma*-infected subjects. The most conspicuous amplifying effect of toxoplasmosis was that on borreliosis–Covid interaction. The effect of being diagnosed with borreliosis was the third strongest effect observed in *Toxoplasma*-infected participants (Tau = 0·212), while in *Toxoplasma*-free subjects, no such effect was observed (Tau = 0·010). The same phenomenon has previously been observed for interaction between toxoplasmosis, borreliosis, and depression.41 A cross-sectional study showed that borreliosis has a relatively strong effect on reported depression but was observed only in *Toxoplasma*-infected participants. Generally, latent toxoplasmosis seems to make human hosts more prone to a wide spectrum of adverse influences, including genetic factors and pathogens.

One can only speculate about the mechanism by which toxoplasmosis influences the risk of SARS-CoV-2 infection and severity of the course of Covid disease, but it is likely that immunomodulation and immunosuppression associated with *Toxoplasma* infection probably play an important role in it. 14-17 Toxoplasmosis has a strong effect on the concentration of various cytokines, especially IL-10, IL-5, IL-6, and TGF-b. Most studies investigating this issue were performed on laboratory animals and might reflect changes associated with acute or post-acute toxoplasmosis rather than its latent stage, but some studies suggest the existence of specific changes in immunity in humans. It has been shown that many parameters of people with latent toxoplasmosis differ from those of *Toxoplasma*-free subjects^18^; for example, seropositive women have increased levels of IL-5 and IL-6, and especially (fivefold) IL-12 in comparison to seronegative controls.^42^ In immunology outpatients, *Toxoplasma*- infected women had increased, and men decreased, counts of leukocytes, NK-cells, and monocytes in comparison to corresponding *Toxoplasma*-free controls.^18^ A cross-sectional study performed on Czech population had shown that *Toxoplasma*-infected subjects more frequently report immune disorders, especially immunodeficiency.^11^

### Limitations and strengths of the study

The main limitation of the present study is that the participants were self-selected and probably do not represent a typical Czech population. Many were recruited via the Facebook site of the main investigator. They are therefore mostly people interested in biological sciences, evolutionary psychology, and quite possibly also cats as pets. On the other hand, they also invited their Facebook friends to participate in the study by pressing the share or like buttons at the end of the questionnaire. Our previous results had shown that the composition of participants of similar internet studies was highly similar, if not identical, to representative internet population with respect to the prevalence of 24 neuropsychiatric disorders^43^ or religiosity.^44^ It is important to emphasise that the informed consent form, as well as the text used for recruitment of participants, mentioned only ‘factors that might influence the risk of infection and the course of Covid disease’, that is, it made no reference to toxoplasmosis.

Toxoplasmosis status was self-reported by respondents, which may be viewed as a drawback. On the other hand, it has previously been demonstrated that information on toxoplasmosis status provided by 3,827 participants of another internet study nearly perfectly (99·5%) corresponded to information in our files on subjects tested for toxoplasmosis in our lab.^45^ Still, about 60% of male and 70% of female respondents recruited via the Facebook-based snowball method were tested for toxoplasmosis elsewhere, mostly in relation to their health problems (49·4% of men) or pregnancy (37·6% of women).^36^ It is possible that some subjects misreported their *Toxoplasma* status. Similarly, some respondents who were *Toxoplasma*-negative during their serological test may have acquired the infection in the meantime. It must be emphasised, however, that presence of misdiagnosed subjects in the population can result in Type 2, not a Type 1 error, that is, it can increase the risk of failure to detect existing effects but not the risk of detecting non-existing effects.

The strength of the study was the large number of participants and the fact that it was preregistered before the start of data collection. Technically, it was thus designed as a cross-sectional study. On the other hand, Covid-19 is a new disease and infected Czech participants probably acquired the infection in autumn 2020, i.e., after being diagnosed with toxoplasmosis. The study thus in fact had the nature of a prospective case-control study and can say at least something about the causal relation between toxoplasmosis and Covid.

## Conclusions

The main result of the present study is the identification of a new risk factor for SARS-CoV-2 infection and severe course of Covid-19. The factor, latent toxoplasmosis, seems to have a stronger effect than most other factors known to affect the risks of Covid-19. Moreover, it seems to enhance the negative effects of some other adverse factors, some of which had not been suspected of having any impact on the course of Covid, such as borreliosis and keeping a cat. The effect of toxoplasmosis is probably rather nonspecific, akin to what has been observed for other diseases and disorders. It is probably related to the observed changes in the immune system of *Toxoplasma* hosts. It can only be speculated whether the effects, e.g. the highly increased level of immunosuppressive cytokine IL-10, are just a side-effect of a latent lifelong infection or part of *Toxoplasma*’s biological adaptations aimed at surviving the attacks of host’s immune system. Latent toxoplasmosis affects about one-third of population in both the developed and developing world. The adverse effects of this zoonosis on public health are therefore probably not negligible.

## Data Availability

The datasets generated and/or analysed during the current study are available in Figshare repository. 10.6084/m9.figshare.14559993.

https://10.6084/m9.figshare.14559993

## Acknowledgements

I would like to thank Anna Pilátová for her useful comments and help with preparing the final version of the article. This work was supported by Czech Science Foundation (grant No. 18-13692S) and Charles University (Research Centre program No. 204056).

## Authors’ contributions

not applicable.

## Conflict of interest statements

The author has no conflict of interests.

## Role of funding source

None.

## Ethics committee approval

The study was approved by the Institutional Review Board of the Faculty of Science, Charles University (Komise pro práci s lidmi a lidským materiálem Přírodovědecké Fakulty Univerzity Karlovy) — No. 2020/25.

## Notes

### Competing Interest Statement

The authors have declared no competing interest.

### Clinical Protocols

https://doi.org/10.17605/OSF.IO/VWXJE

### Author Declarations

The study was approved by the Institutional Review Board of the Faculty of Science, Charles University (Komise pro praci s lidmi a lidskym materialem Prirodovedecke Fakulty Univerzity Karlovy) No. 2020/25.

## References

1. Tenter AM, Heckeroth AR, Weiss LM. Toxoplasma gondii: from animals to humans. Int J Parasitol 2000; 30(12-13): 1217-58.

2. El-Saadi O, Welham J, Saha S, et al. The incidence and prevalence of schizophrenia: Preliminary results from a systematic review. Schizophr Res 2002; 53(3): 32-.

3. Torrey EF, Bartko JJ, Lun ZR, Yolken RH. Antibodies to Toxoplasma gondii in patients with schizophrenia: A meta-analysis. Schizophr Bull 2007; 33: 729–36.

4. Torrey EF, Bartko JJ, Yolken RH. Toxoplasma gondii and other risk factors for schizophrenia: An update. Schizophr Bull 2012; 38(3): 642–7.

5. Flegr J, Horáček J. Toxoplasma-infected subjects report an obsessive-compulsive disorder diagnosis more often and score higher in obsessive-compulsive inventory. Eur Psychiat 2017; 40: 82– 7.

6. Sutterland AL, Fond G, Kuin A, et al. Beyond the association. Toxoplasma gondii in schizophrenia, bipolar disorder, and addiction: systematic review and meta-analysis. Acta Psychiatr Scand 2015; 132(3): 161–79.

7. Lindová J, Kuběna AA, Šturcová A, et al. Pattern of money allocation in experimental games supports the stress hypothesis of gender differences in Toxoplasma gondii-induced behavioural changes. Folia Parasitol 2010; 57: 136–42.

8. Lindová J, Novotná M, Havlíček J, et al. Gender differences in behavioural changes induced by latent toxoplasmosis. Int J Parasitol 2006; 36: 1485–92.

9. Flegr J, Zitkova S, Kodym P, Frynta D. Induction of changes in human behaviour by the parasitic protozoan Toxoplasma gondii. Parasitology 1996; 113: 49–54.

10. Flegr J. Influence of latent Toxoplasma infection on human personality, physiology and morphology: pros and cons of the Toxoplasma-human model in studying the manipulation hypothesis. J Exp Biol 2013; 216(1): 127–33.

11. Flegr J, Escudero DQ. Impaired health status and increased incidence of diseases in Toxoplasma-seropositive subjects - an explorative cross-sectional study. Parasitology 2016; 143(14): 1974–89.

12. Flegr J, Prandota J, Sovickova M, Israili ZH. Toxoplasmosis - A global threat. Correlation of latent toxoplasmosis with specific disease burden in a set of 88 countries. PLoS ONE 2014; 9(3).

13. Havlíček J, Gašová Z, Smith AP, Zvára K, Flegr J. Decrease of psychomotor performance in subjects with latent ‘asymptomatic’ toxoplasmosis. Parasitology 2001; 122: 515–20.

14. Neyer LE, Grunig G, Fort M, Remington JS, Rennick D, Hunter CA. Role of interleukin-10 in regulation of T-cell-dependent and T-cell-independent mechanisms of resistance to Toxoplasma gondii. Infect Immun 1997; 65(5):1675–82.

15. Kaňková Š, Holáň V, Zajícová A, Kodym P, Flegr J. Modulation of immunity in mice with latent toxoplasmosis - the experimental support for the immunosupression hypothesis of Toxoplasma-induced changes in reproduction of mice and humans. Parasitol Res 2010; 107: 1421–7.

16. Buzoni-Gatel D, Dubremetz JF, Werts C. Molecular cross talk between Toxoplasma gondii and the host immune system. M S-Medecine Sciences 2008; 24: 191–6.

17. Fenoy IM, Chiurazzi R, Sanchez VR, et al. Toxoplasma gondii infection induces suppression in a mouse model of allergic airway inflammation. PLoS ONE 2012; 7(8).

18. Flegr J, Stříž I. Potential immunomodulatory effects of latent toxoplasmosis in humans. BMC Infect Dis 2011; 11: 274.

19. Kankova S, Flegr J, Calda P. An elevated blood glucose level and increased incidence of gestational diabetes mellitus in pregnant women with latent toxoplasmosis. Folia Parasitol 2015; 62.

20. R Core Team. R: A language and environment for statistical computing. R Foundation for Statistical Computing. Vienna. Austria; 2018.

21. Kim S. ppcor: An R package for a fast calculation to semi-partial correlation coefficients. Commun Stat Appl Methods 2015; 22(6): 665–74.

22. Benjamini Y, Hochberg Y. Controlling the false discovery rate: A practical and powerful approach to multiple testing. J Roy Stat Soc B Met 1995; 57(1): 289–300.

23. Novotná M, Havlíček J, Smith AP, et al. Toxoplasma and reaction time: Role of toxoplasmosis in the origin, preservation and geographical distribution of Rh blood group polymorphism. Parasitology 2008; 135: 1253–61.

24. Flegr J, Novotná M, Fialová A, Kolbeková P, Gašová Z. The influence of RhD phenotype on toxoplasmosis- and age-associated changes in personality profile of blood donors. Folia Parasitol 2010; 57: 143–50.

25. Flegr J, Preiss M, Klose J. Toxoplasmosis-associated difference in intelligence and personality in men depends on their Rhesus blood group but not ABO blood group. PLoS ONE 2013; 8(4).

26. Waynforth D. Life-history theory, chronic childhood illness and the timing of first reproduction in a British birth cohort. Proceedings Biological sciences 2012; 279(1740): 2998–3002.

27. Flegr J, Hrušková M, Hodný Z, Novotná M, Hanušová J. Body height, body mass index, waisthip ratio, fluctuating asymmetry and second to fourth digit ratio in subjects with latent toxoplasmosis. Parasitology 2005; 130: 621–8.

28. Hodkova H, Kolbekova P, Skallova A, Lindova J, Flegr J. Higher perceived dominance in Toxoplasma infected men - a new evidence for role of increased level of testosterone in toxoplasmosis-associated changes in human behavior. Neuroendocrinol Lett 2007; 28(2): 110–4.

29. Flegr J, Lindová J, Kodym P. Sex-dependent toxoplasmosis-associated differences in testosterone concentration in humans. Parasitology 2008; 135: 427–31.

30. Kaňková Š, Kodym P, Flegr J. Direct evidence of Toxoplasma-induced changes in serum testosterone in mice. Exp Parasitol 2011; 128: 181–3.

31. Lim A, Kumar V, Hari Dass SA, Vyas A. Toxoplasma gondii infection enhances testicular steroidogenesis in rats. Mol Ecol 2013; 22(1): 102–10.

32. Tan D, Vyas A. Toxoplasma gondii infection and testosterone congruently increase tolerance of male rats for risk of reward forfeiture. Horm Behav 2016; 79: 37–44.

33. Leroy EM, Gouilh MA, Brugere-Picoux J. The risk of SARS-CoV-2 transmission to pets and other wild and domestic animals strongly mandates a one-health strategy to control the COVID-19 pandemic. One Health 2020; 10.

34. Newman A, Smith D, Ghai RR, et al. First reported cases of SARS-CoV-2 infection in companion animals - New York, March-April 2020. MMWR-Morb Mortal Wkly Rep 2020; 69(23):710–3.

35. Hosie MJ, Hofmann-Lehmann R, Hartmann K, et al. Anthropogenic infection of cats during the 2020 COVID-19 pandemic. Viruses 2021; 13(2).

36. Flegr J, Preiss M. Friends with malefit. The effects of keeping dogs and cats, sustaining animal-related injuries and Toxoplasma infection on health and quality of life. PLoS ONE 2019; 14(11).

37. Hanauer DA, Ramakrishnan N, Seyfried LS. Describing the relationship between cat bites and human depression using data from an electronic health record. PLoS ONE 2013; 8(8): e70585.

38. Flegr J, Hodny Z. Cat scratches, not bites, are associated with unipolar depression - cross-sectional study. Parasit Vectors 2016; 9.

39. Flegr J, Vedralova M. Specificity and nature of the associations of twenty-four neuropsychiatric disorders with contacts with cats and dogs. Schizophr Res 2017; 189: 219–20.

40. Flegr J, Preiss, M., Balatova, P. Depressiveness and neuroticism in Bartonella seropositive and seronegative subjects-Preregistered case-controls study. Front Psychiatry 2018; 9.

41. Flegr J, Horáček J. Toxoplasmosis, but not borreliosis, is associated with psychiatric disorders and symptoms. Schizophr Res 2018; 197: 603–4.

42. Matowicka-Karna J, Dymicka-Piekarska V, Kemona H. Does Toxoplasma gondii infection affect the levels of IgE and cytokines (IL-5, IL-6, IL-10, IL-12, and TNF-alpha)? Clin Dev Immunol 2009; 374696: 374696.

43. Flegr J, Horáček J. Negative effects of latent toxoplasmosis on mental health. Front Psychiatry 2020; 10.

44. Flegr J, Kuba R, Kopecký R. Rhesus-minus phenotype as a predictor of sexual desire and behavior, wellbeing, mental health, and fecundity. PLoS ONE 2020; 15(7): e0236134.

45. Flegr J. Predictors of Toxoplasma gondii infection in Czech and Slovak populations: the possible role of cat-related injuries and risky sexual behavior in the parasite transmission. Epidemiol Infect 2017; 145(7): 1351–62.

